# The potential impact of the spreading of highly transmissible Omicron variant XBB.1.5 and JN.1 on the evolution of SARS-CoV-2

**DOI:** 10.1101/2024.04.20.24306131

**Authors:** Buqing Yi

## Abstract

In spite of three-year pandemic time caused by Covid-19, the trend and impact of the evolution of SARS-CoV-2 remain unclear. For newly emerging variants, it is still hard to predict how possibly they could impact the pandemic/endemic course. The spreading of the highly transmissible variant XBB.1.5 and JN.1 has brought many questions. Could the high transmission ability of such variant lead to a different global spreading pattern compared to other previously existing variants of concern (VOCs)? How that may possibly affect the further evolution of SARS-CoV-2? This study aimed to explore the evolution course of SARS-CoV-2 variants, with focus on VOCs. The investigation was carried out through integration of multiple virus genomic epidemiology approaches. Phylogeny and phylodynamic analyses were used to investigate dynamic changes of virus evolution course. Through this study, an overview check of the evolution events of SARS-CoV-2 VOCs, including the alpha variant, delta variant and omicron variant, has been performed, and the further course of evolution has been predicted, especially taking the potential impact of the spreading of XBB.1.5 and JN.1 into consideration. Furthermore, the early spreading pattern of XBB.1.5 was compared with the early spreading patterns of BQ.1.1 to investigate the impact of evolution on virus spreading pattern. The information acquired through this study provided insights into the effect of SARS-CoV-2 evolution on spreading pattern and the potential impact of the spreading of XBB.1.5 and JN.1 on the further evolution of SARS-CoV-2.

## 1. Introduction

Covid-19 led to three-year pandemic time and caused lots of health problems as well as significant economic loss. However, despite huge efforts being made, till now it is still hard to predict the trend and impact of the evolution of SARS-CoV-2 regarding the public health implications. In light of high levels of population immunity in many settings and country differences in the immune landscape, the evolution of SARS-CoV-2 has been under close monitoring globally. In particular, the emergence of highly transmissible variant XBB.1.5 and other related XBB sub-lineages (indicated as XBB*) has brought many questions. The newly emerging variants of interest (VOIs) JN.1 further emphasizes the importance of investigating the trend and impact of the SARS-CoV-2 evolution.

### 1.1. The evolution of omicron variant

The Omicron variant evolved quickly from the aspect of SARS-CoV-2 evolution, with the first variant BA.1 carrying more than 60 mutations (Cui et al., 2022), and in general showed decreased hospitalization rates and less severe disease in patients (Maslo et al., 2022). However, it is unclear how the Omicron variants reduce pathogenicity, and it is also unknown if the reduced pathogenicity promotes transmission. In Europe, BA.1 was quickly replaced by BA.2, which was followed by BA.5. BQ.1* is a sublineage of BA.5, which carries spike mutations in some key antigenic sites, including K444T and N460K. In addition to these mutations, the sublineage BQ.1.1 carries an additional spike mutation in a key antigenic site (i.e. R346T). It is likely that these additional mutations have conferred an immune escape advantage over other circulating Omicron sublineages, and therefore a possible higher reinfection risk.

### 1.2. The emergence of XBB lineage

BQ.1* displayed a significant growth advantage over other circulating Omicron sub-lineages in many settings and became one of the predominant Omicron variants in many countries in late 2022. However, in early 2023, it was largely outcompeted by XBB.1.5. The XBB* lineage was first identified by public health authorities in India during summer 2022, which arose as a result of co-infection of two Omicron subvariants: BA.2.10.1 and BA.2.75 (Singh, Sharma, Shaw, Bhargava, & Negi, 2023). As of epidemiological week 40 (3 to 9 October) in 2022, from the sequences submitted to GISAID, XBB* had a global prevalence of 1.3% and it had been detected in 35 countries. As of epidemiological week 40 (1 to 7 October) in 2023, from the sequences submitted to GISAID, XBB* had a global prevalence of 95.7% and it had been detected in most countries around the world. XBB.1.5 is a direct descendent of the original XBB variant. It was first detected in USA in October 2022, then quickly spread to Europe.

It is unclear why XBB.1.5 displayed growth advantage over BQ.1.1. In late 2022 and early 2023, XBB*, especially XBB.1.5, XBB.1.16, XBB.1.9, displayed growth advantage over other co-circulating variants in many countries. Based on currently available evidence, it appears that the overall phenotype of XBB* does not diverge sufficiently from other Omicron lineages with additional immune escape mutations (such as BQ.1.1). Till December 2024, the Omicron variant of concern (mainly BA.2.86 & JN.1) remained the dominant variant circulating globally, accounting for almost all sequences reported to GISAID. Based on currently available information, the vast genetic diversity of Omicron sub-lineages has been mainly manifested by differences in immune escape potential, instead of clinical outcomes.

### 1.3. The relationship between virus evolution and epidemiological events

In spite of many efforts being made to investigate the evolution of SARS-CoV-2, till now it is still unclear what can be the future trend of SARS-CoV-2 evolution. In particular, the emergence of XBB.1.5 and BA.2.86 & JN.1 which harbor many mutations raises the need to further explore the trend and impact of SARS-CoV-2 evolution. In this paper, we would focus on the XBB.1.5 involved pandemic course in Europe to inquire information regarding virus evolution and spreading pattern, which might be able to deepen our understanding about the relationship between virus evolution and epidemiological events, and help prediction of future evolution trend and/or the potential risk of any new variant causing infection waves.

## 2. Methods

### 2.1. Establishment of genome sequence data set for genomic epidemiology investigation

We used full-length SARS-CoV-2 sequences downloaded from GISAID (Shu & McCauley, 2017) to build up genome sequence data set for epidemiology investigation. We performed quality check and filtered out low-quality sequences that met any of the following criteria: 1) sequences with less than 90% genome coverage; 2) genomes with too many private mutations (defined as having >24 mutations relative to the closest sequence in the reference tree); 3) genomes with more than ten ambiguous bases; and 4) genomes with mutation clusters, defined as 6 or more private differences within a 100-nucleotide window. These are the standard quality assessment parameters utilized in NextClade (https://clades.nextstrain.org) (Aksamentov, Roemer, Hodcroft, & Neher, 2021).

### 2.2. Lineage classification

We used the dynamic lineage classification method through the Phylogenetic Assignment of Named Global Outbreak Lineages (PANGOLIN) software suite (https://github.com/hCoV-2019/pangolin) (Rambaut et al., 2020). This is intended for identifying the most epidemiologically important lineages of SARS-CoV-2 at the time of analysis (O’Toole et al., 2021).

### 2.3. Phylogenetic and phylogeographical analyses of SARS-CoV-2

Phylogenetic analysis and phylogeographical analyses were carried out to infer the transmission routes of BQ.1.1 and XBB.1.5 in Europe (Dellicour, Rose, & Pybus, 2016) and to track evolution event with a custom build of the SARS-CoV-2 NextStrain build (https://github.com/nextstrain/ncov) (Hadfield et al., 2018; Stadtmüller et al., 2022; Yi et al., 2024; Yi et al., 2021). The pipeline includes several Python scripts that manage the analysis workflow. Briefly, it allows for the filtering of genomes, the alignment of genomes in NextClade (https://clades.nextstrain.org) (Aksamentov et al., 2021), phylogenetic tree inference in IQ-Tree (Minh et al., 2020; Tony-Odigie, Dalpke, Boutin, & Yi, 2024; Yi & Dalpke, 2021), tree dating (Sagulenko, Puller, & Neher, 2018; Yi, Bumbarger, & Sommer, 2009) and ancestral state construction and annotation. The phylogeny analysis is rooted by *Wuhan*-Hu-1/2019 (GISAID Accession ID: EPI_ISL_402125). To infer the transmission routes of BQ.1.1 and XBB.1.5 in Europe, only samples fulfilling these criteria on GISAID were included in the analysis: 1. With complete sample collection dates; 2. With a complete sequence (>29,000nt) and less than 5% Ns.

### 2.4. Evaluation of phylogenetic distances among the representative VOCs and VOIs

The SRAS-CoV-2 VOC Alpha, Beta, Gamma, Lambda, Mu, Delta, BA.1, BA.2, BA.3, BA.4, BA.5, BF.7, BQ.1.1, and VOI XBB.1.5*, BA.2.86* XBB.1.16*, EG.5*, are included in the analysis. For each VOC or VOI, ten samples (five from the early period and five from the late period of its circulation) were randomly chosen and used for the analysis. Phylogenic distances (shown as values of divergence from the B variant) were evaluated by running the SARS-CoV-2 NextStrain build (https://github.com/nextstrain/ncov) (Hadfield et al., 2018; Stadtmüller et al., 2022; Yi et al., 2021).

### 2.5. Relative growth advantage

We analyzed SARS-CoV-2 sequences from Europe that were uploaded to GISAID with complete sample collection dates from September 1^st^ to October 15^th^ 2022 (for the analysis of BQ.1.1*) or from December 1^st^ 2022 to January 15^th^ 2023 (for the analysis of XBB.1.5*) or from October 1^st^ 2023 to November 15^th^ 2023 (for the analysis of JN.1*). A logistic regression model was used to estimate the relative growth advantage of certain variant compared to co-circulating variants as previously reported (Campbell et al., 2021; Chen, Nadeau, Yared, et al., 2021; Chen, Nadeau, Topolsky, et al., 2021; Davies et al., 2021). The model assumes that the increase or decrease of the proportion of a variant follows a logistic function, which is fit to the data by optimizing the maximum likelihood to obtain the logistic growth rate in units per day. Based on that, an estimate of the growth advantage per generation is obtained (assuming the growth advantage arising from a combination of intrinsic transmission advantage, immune evasion, and a prolonged infectious period (Althaus et al., 2021), and the relative growth advantage per week (in percentage; 0% means equal growth) is reported. The relative growth advantage estimate reflects the advantage compared to co-circulating variants in the selected region and time frame. The analyses were primarily performed with RStudio v1.3.1093 with multiple R software (Muhandes et al., 2021; Tony-Odigie, Wilke, Boutin, Dalpke, & Yi, 2022; Yi, Matzel, et al., 2015; Yi, Rykova, et al., 2015; Yi, Titze, et al., 2015), e.g. tidyverse, ggplot.

## 3. Results

### 3.1. Unique mutations detected in XBB.1.5* or BA.2.86/JN.1

XBB.1.5 is like a collection of mutations present in the previous VOCs. It contains mutations present in many other VOCs (Harvey et al., 2021), not only mutations present in Omicron, but also in other previously detected VOCs, such as Alpha (e.g. V143-; Y144-) (McCarthy et al., 2021). If each mutation has certain impact on transmission or pathogenicity of the virus, then in XBB.1.5 an accumulative effect can be assumed. Only a few mutations being frequently detected in other VOCs are absent in XBB.1.5, e.g. H69-; V70-; L452R (Zhang et al., 2022). XBB.1.5 and several co-circulating variants BQ.1.1, BF.7, XAY.1.1, BF.11, CQ.2, BN.1.3, BA.4.6 share a few unique mutations, e.g. R346T; N460K, which are absent in the previously circulating VOCs including BA.4/BA.5, suggesting potential functional importance and a fitness advantage of these mutations.

**Table 1:**
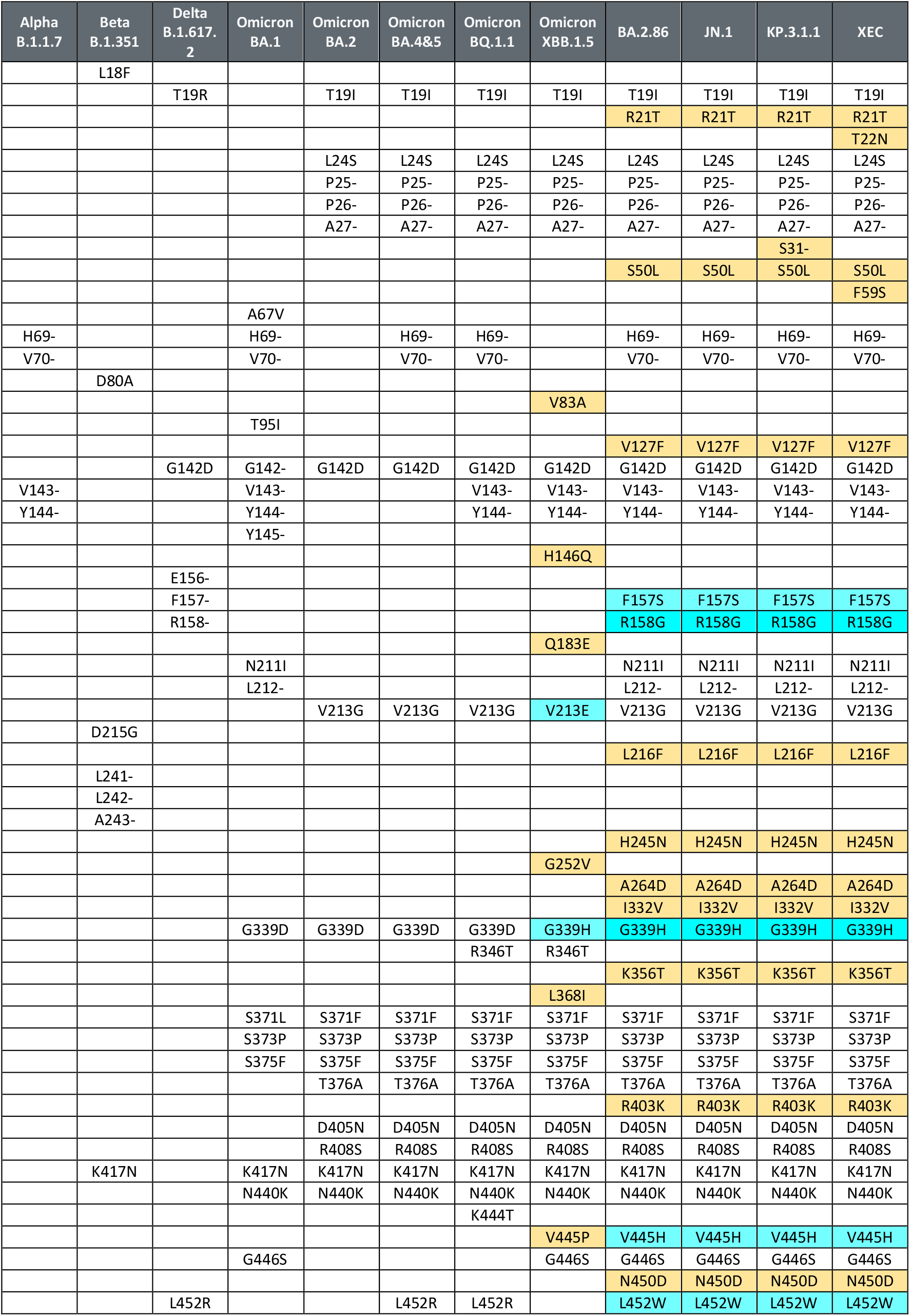
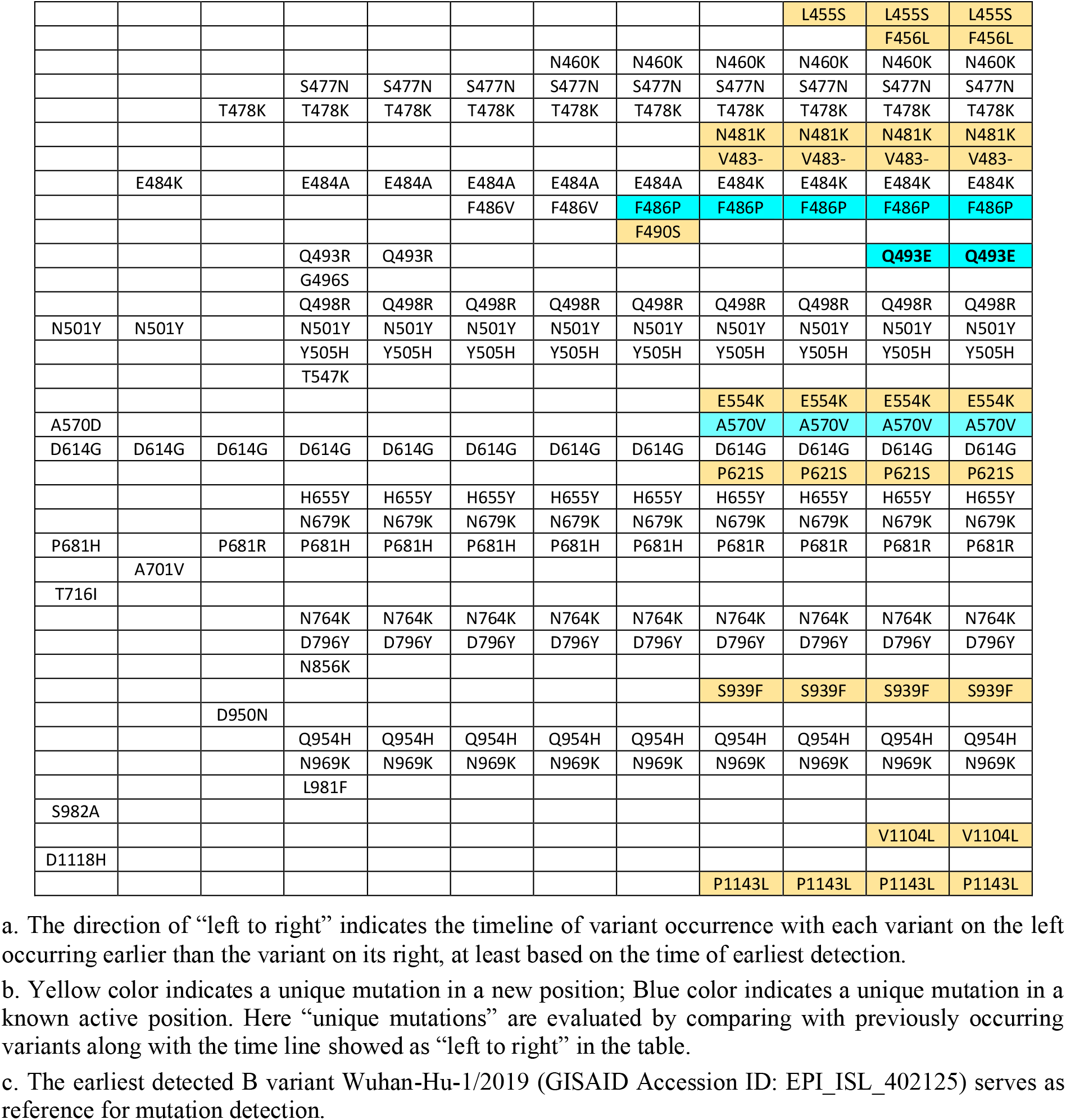
Spike mutation comparison among representative VOCs/VOIs.

Mutations uniquely detected in XBB.1.5 have two types (some also present in other XBB sub-lineages): a. In known active sites (the positions where mutations have been detected in other VOCs) but unique mutations, e.g. G339H instead of G339D; V213E instead of V213G; F486P instead of F486V. b. In positions where mutations have been rarely detected previously, e.g. V83A, H146Q and Q183E.

Compared to XBB.1.5*, which is manifested as a collection of previously occurred mutations, BA.2.86* displays an evolution jump with many new mutations (>20) occurring in this variant. Interestingly, none of the unique mutations in XBB.1.5* (in positions without previously detected mutation) was detected in BA.2.86*. On the background of BA.2.86, JN.1* has one extra mutation L455S, which was rarely detected in other variants. JN.1 further evolved to various subvariants, such as KP.3, which carries one extra mutation Q493E compared to JN.1. KP.3 was able to further evolve to multiple subvariants. In particular, KP.3.1.1 outcompeted KP.3 to become the predominant strain globally. It is noteworthy that compared to KP.3, KP.3.1.1 only carries one extra S31 deletion (Feng et al., 2024). Another JN.1 subvariant, XEC, is rapidly expanding globally and will possibly become the next predominant strain. XEC is actually one recombinant variant of KS.1.1 and KP.3.3 (Li et al., 2024), showing a high similarity with KP.3 with only two additional spike mutations F59S and T22N.

### 3.2. Phylogenetic distances of representative VOCs/VOIs

We evaluated phylogenetic distances among representative VOCs and VOIs by “divergence” based on the relevant algorithm applied in Nextstrain. In Figure 1, the values of divergence of each VOC or VOI from the earliest detected B variant are displayed. For B variant itself, the value of divergence is zero, indicating no divergence. In the pre-Omicron time, the Alpha, Delta, Gamma, Mu and Lambda display a similar divergence of around 40. The Omicron variants showed a jump of evolution, with BA.1, BA.2 and BA.3 exhibiting a divergence of around 60. For BA.4, BA.5 and its sub-lineages BQ.1.1 as well as BF.7, the divergence increased to around 80. For XBB* lineages, such as XBB.1.5*, XBB.1.16*, EG.5*, the divergence further escalated to between 100 and 120. Strikingly, the divergence of BA.2.86*/JN.1* lineages shot up to around 180, which is three times higher than that of the early Omicron variants BA.1, and almost two times higher than that of XBB.1.5, suggesting a special speed-up evolutionary event taking place in BA.2.86*/JN.1*. Previously the Omicron variants with a high divergence from the pre-Omicron variants displayed pathogenicity changes resulting in milder symptoms (Menni et al., 2022). Till December 2024, it was still unclear if the JN.1 variant causes different symptoms, but its high divergence from other previous variants suggests possible transmissibility and/or pathogenicity changes. KP.3.1.1 and XEC are two subvariants of JN.1 spreading globally. The divergence of these two variants slightly further increased to between 190 and 200, showing a limited evolution from BA.2.86*/JN.1*.

**Figure 1:**
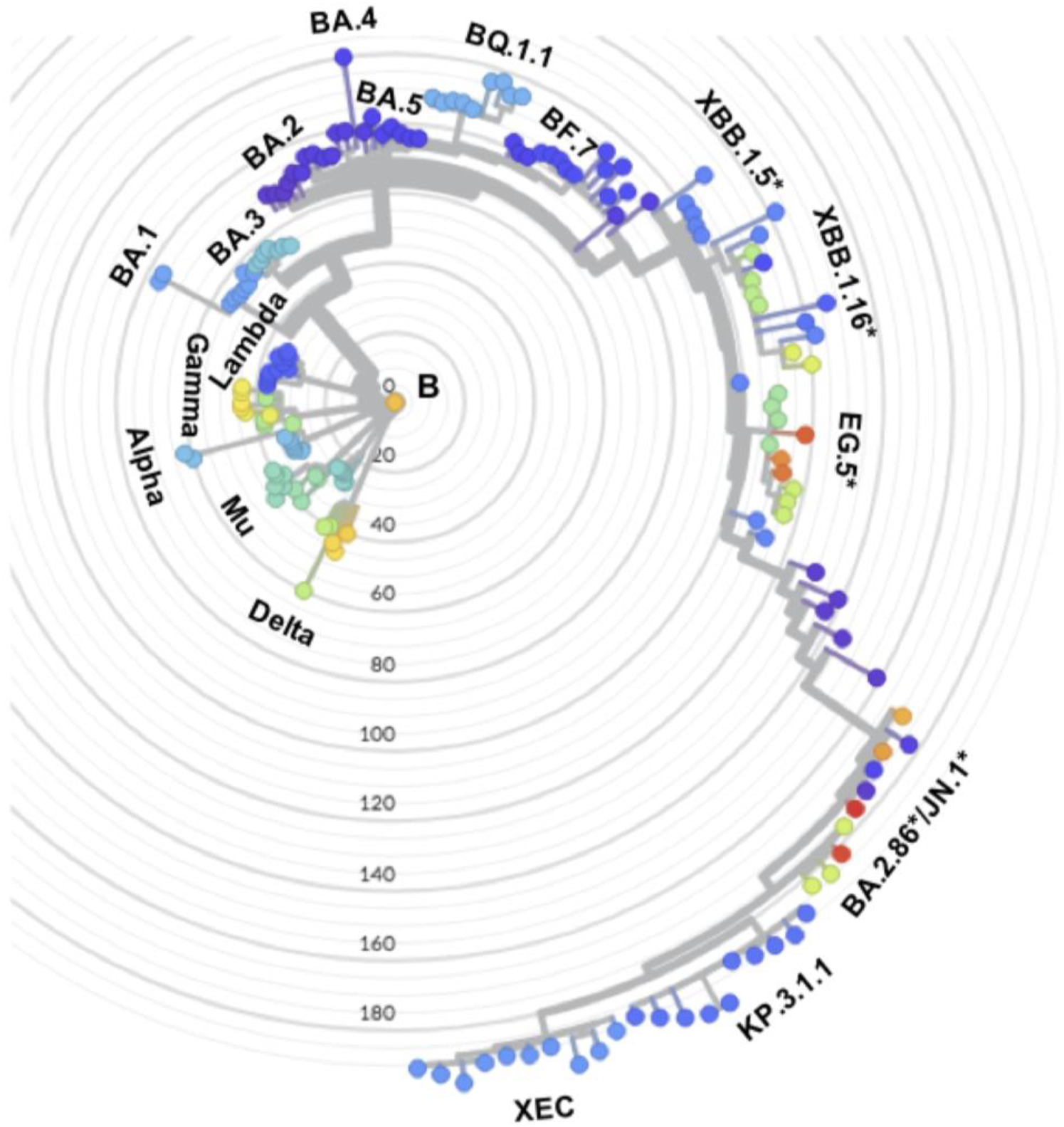
Phylogenetic distances among representative VOCs and VOIs. Each color represents one specific lineage. The names of VOCs and VOIs are displayed next to the corresponding lineage groups, which usually include multiple sub-lineages. Numbers indicate phylogenetic distance of “divergence” as calculated through the algorithm in Nextstrain. B is the earliest detected SARS-CoV-2 variant serving as reference for calculating phylogenetic distance. For each VOC or VOI, ten randomly chosen samples are used in the analysis. The analysis is based on data status on 2024-11-30.

### 3.3. Comparison of growth advantage of JN.1, XBB.1.5, BQ.1.1 and BA.1 over their respective co-circulating variants during their early spreading periods

In late 2021, the Omicron variant BA.1 was able to outcompete the Delta variants and became the dominant variant (Yi et al., 2022). Till late 2024, the Omicron variants were still the dominant SARS-Cov-2 variants, but with a complicated evolution course. In late 2023, JN.1 was able to outcompete XBB* variants e.g. XBB.1.5* and EG.5*, and became the predominant variant in Europe. Before the emergence of XBB.1.5 in late 2022 in Europe, BQ.1.1 was one of the major dominant Omicron variants. Among these variants, JN.1* and XBB.1.5* display huge genetic differences, while XBB.1.5* and BQ.1.1 share many mutations. They were all able to outcompete other co-circulating variants via a growth advantage. To understand the impact of evolution on spreading patterns, here we first analyzed the relative growth advantage of these variants over their respective co-circulating variants during their early spreading periods.

The relative growth advantage estimate reflects the advantage compared to co-circulating variants in the selected region and time frame. In this analysis, we focused on the period of the first one and half month starting from the time when community transmission of each variant became detectable in Europe (see details below for the time window identification). Considering the sequencing efforts are roughly comparable between 2021 to early 2023, the results in this period can be reliably compared to each other as well, but results from late 2023 might be less comparable owing to the reduction of sequencing efforts.

During their early spreading period in Europe, BA.1 (between Nov. 15^th^ and Dec. 31^st^, 2021) and JN.1 (between Oct. 1^st^ and Nov. 15^th,^ 2023) both showed a remarkable relative growth advantage of 116% and 93%, respectively (Figure 3A&B). BQ.1.1 (between Sep. 1^st^ and Oct. 30^th^, 2022) displayed a relative growth advantage of around 70 % compared to co-circulating variants (mainly other BA.5 sub-lineages, e.g. BA.5 and BF.7), and XBB.1.5 (between Dec. 1^st^ 2022 till Jan. 15^th^ 2023) displayed a relative growth advantage of 80% compared to co-circulating variants (mainly BA.5 sub-lineages, e.g. BF.7 and BQ.1.1) (Figure 2). This means, JN.1 has a growth advantage over XBB.1.5, and XBB.1.5 has a growth advantage over BQ.1.1.

**Figure 2:**
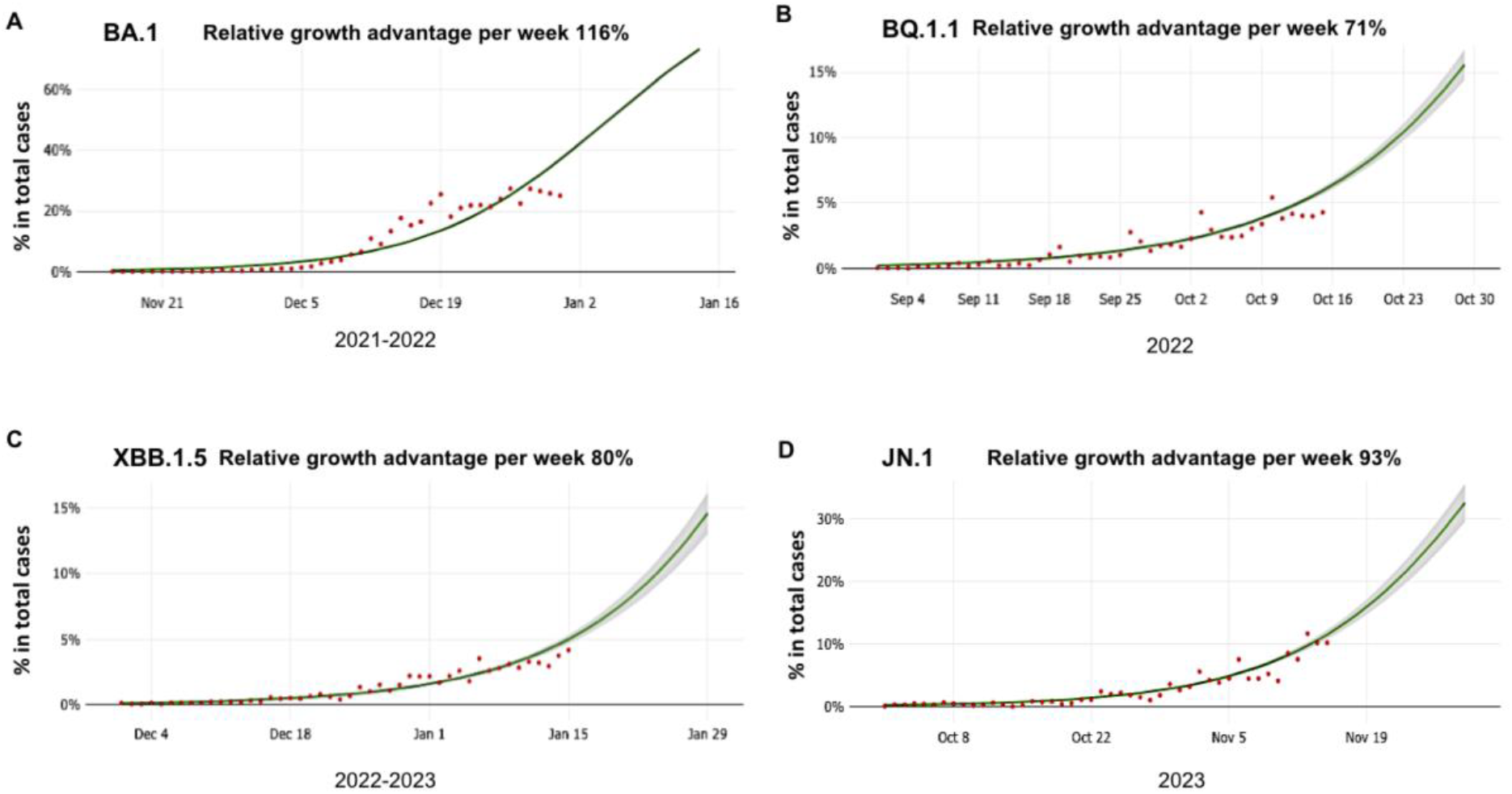
Growth of BA.1, BQ.1.1, XBB.1.5 and JN.1 during their early spreading period in Europe. Model fits are based on a logistic regression. Dots represent the daily proportions of variants. The relative growth advantage per week (in percentage; 0% means equal growth) is reported. The shaded areas correspond to the 95% CIs of the model estimates. The BA.1, BQ.1.1, XBB.1.5 and JN.1 all spread quickly in Europe with a relative growth advantage of around 116%, 71%, 80% and 93%, compared to their co-circulating variants, respectively.

**Figure 3:**
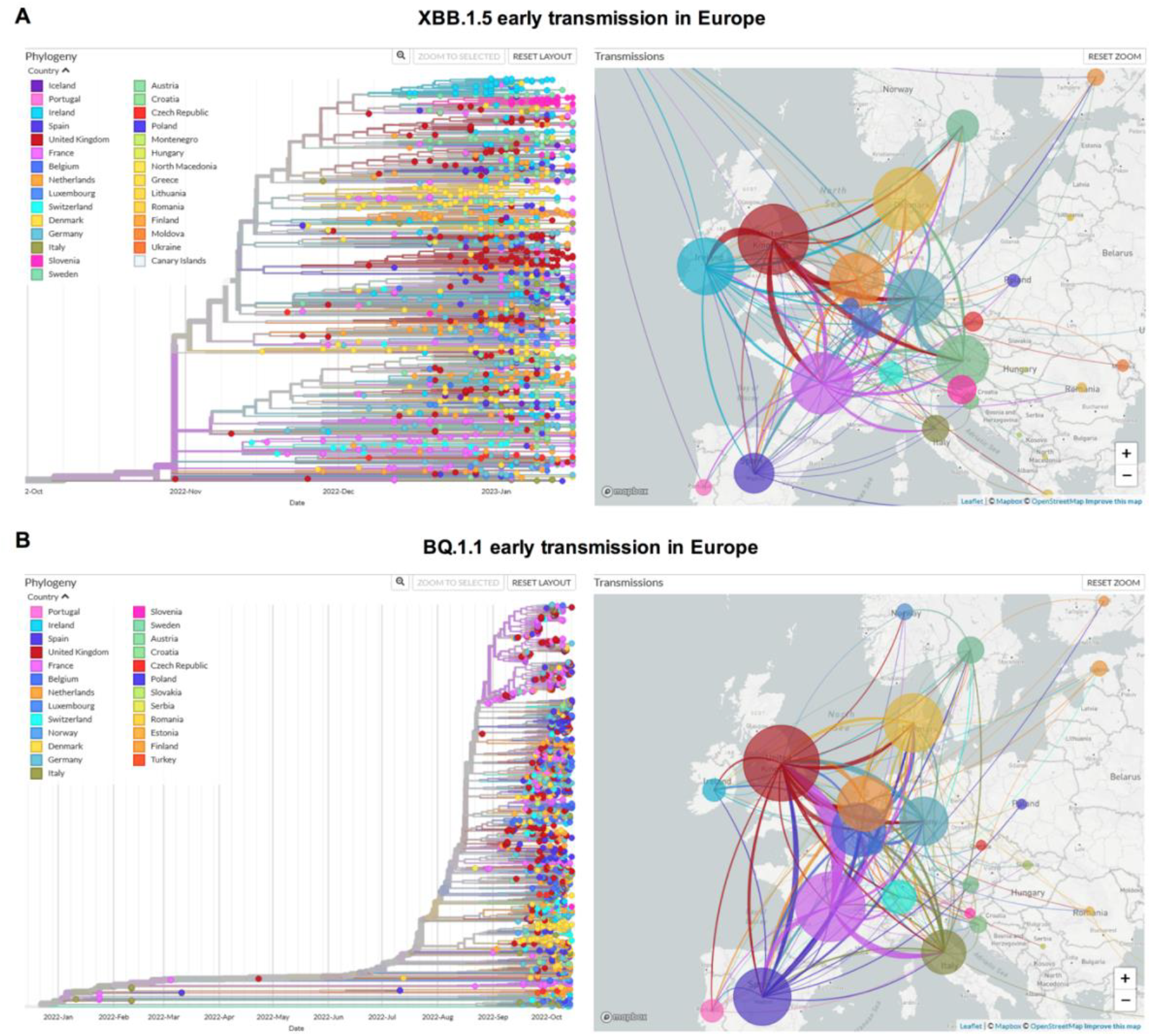
Early transmission routes of XBB.1.5 and BQ.1.1 in Europe inferred based on phylogeny analysis. The phylogeny analysis was based on the earliest 2000-3000 samples detected in Europe for each variant. The size of the circle represents the number of genomes from the XBB.1.5 (A) or from the BQ.1.1 (B) in each country. The line colors correspond to the exporting locations. **A**. Left: Phylogeny tree of XBB.1.5 collected in Europe till 15.01.2023, with branch length representing time; Right: Estimated early transmission events of XBB.1.5 in Europe mainly taking place between end of 2022 and early 2023 (till 15.01.2023). **B**. Left: Phylogeny tree of BQ.1.1 collected in Europe till 15.10.2022, with branch length representing time; Right: Estimated early transmission events of BQ.1.1 in Europe mainly taking place between September to early October of 2022 (till 15.10.2022). For both variant, the transmission events displayed here took place in around 45 days.

### 3.4. Comparison of the early spreading of JN.1, XBB.1.5 and BQ.1.1 in Europe

Although XBB.1.5 and BQ.1.1 share many mutations, XBB.1.5 was able to outcompete BQ.1.1 in many places globally including Europe. What is the advantage of XBB.1.5? What are the major mutations in XBB.1.5 that promote the transmission of XBB.1.5 surpassing BQ.1.1? Had the high transmission ability of XBB.1.5 been able to lead to a different global spreading pattern compared to other previously existing variants of concern (VOCs)? Till now, these questions have not been fully addressed yet.

Here we would like to focus on the comparison of early spreading pattern of XBB.1.5 and BQ.1.1 in Europe. The objective of this section is to investigate the impact of evolution on virus spreading pattern. The similar analyses were not applicable for BA.2.86* or JN.1* as many countries stopped or largely decreased virus genomic sequencing in late 2023.

In Europe, the spreading routes of XBB.1.5 pretty much resemble the traffic network in Europe with virus exporting and importing being largely balanced for most countries, and none of the countries was revealed as the prominent exporting location of XBB.1.5 (Figure 3A). This concurs with the known information that XBB.1.5 was first detected in North America in October 2022. Through cross-continent traffic it got spread to Europe since late October on, and then since early December, community transmission of XBB.1.5 has been detected in Europe (Figure 3A). The time lag between the first sample being detected till community transmission becoming visible was only around one month. The countries in which the earliest community transmissions were detected are overlapped with the countries with busiest traffic connection with North America, such as France and the UK. This is a good example of how traffic network may affect virus global spreading.

On the contrary, the early spreading of BQ.1.1 was very different from that of XBB.1.5. In January 2022, BQ.1.1 was already detected in a few European countries, such as Italy and France, but community transmission of BQ.1.1 in Europe was not visible until late August 2022 (Figure 3B). The time lag between the first sample being detected till community transmission becoming visible was around eight months, displaying a big difference from that of XBB.1.5. Part of the reason can be the different source location of these two variants. The source location of XBB.1.5 was North America, which means the earliest spreading of it in Europe was through cross-continent traffic, and the spreading would be constant if there were already a big number of cases in the source country. If the traffic volume with north-America is roughly comparable, most European countries would have imported cases in a similar scale, which means community transmission would take place in most countries at a similar time point (although France and the UK started community transmission slightly earlier than the other countries). This is exactly what has been observed as shown in Figure 3B.

For BQ.1.1, the earliest samples were almost exclusively detected in Europe, suggesting its source location was one of European countries. For newly emerging variant, if it has no significant fitness advantage compared to other co-circulating variant, the case number increase would be very slow at the early period. The other reason can be, the early samples of BQ.1.1 were a few randomly occurred cases, the spreading of which was inhibited somehow through non-pharmaceutical intervention applied in many countries in Europe in early 2022. Later in July, through independent evolution events, BQ.1.1 emerged again in Europe and developed into visible community transmission in late August 2022. In this case, the time lag between the first sample being detected till community transmission becoming visible was around one and half months, which was only slightly longer than that of XBB.1.5.

## 4. Discussion

This study aimed to explore the interplay between SARS-CoV-2 evolution and spreading, with focus on VOCs and VOIs. Through integration of multiple virus genomic epidemiology approaches, we first investigated the evolution course of SARS-CoV-2 variants. Both mutation analysis and phylogenetic distance evaluation have revealed two evolutionary jumps during the SARS-CoV-2 evolution, the first one was from the pre-Omicron variants to the Omicron variant BA.1, and the second one was from the XBB* variants to BA.2.86*/JN.1*. It is noteworthy that in Europe the Omicron variant BA.1 outcompeted the Delta variants with a very high relative growth advantage of 116%, and JN.1 was also able to outcompete XBB* variants with a high relative growth advantage of 93%. In both cases, the evolution course did not follow the step-wise procedure, whereas a dramatic change of many unique mutations took place. Epidemiological investigation of BQ.1.1 (one BA.5 sub-lineage) and XBB.1.5 revealed different spreading patterns in Europe, which might be partially affected by the genetic differences of these two variants, and partially affected by geographical factors. These data suggest genetic changes of SARS-CoV-2 variants may affect epidemiological events, at least to a certain extent.

### 4.1. The impact of mutations in BA.2.86 and XBB.1.5 on functions and pathogenicity

Although currently it remains a challenging task to predict the evolution course of SARS-CoV-2 or to pinpoint potential future dominant variants, many SARS-CoV-2 virus functional analyses have been performed to unveil the impact of virus evolution on functions. It has been shown that SARS-CoV-2 Omicron variants BQ.1.1 and XBB.1* (e.g. XBB.1 or XBB.1.5) both displayed remarkable neutralization escape capability compared to non-Omicron or the early Omicron variants (Miller et al., 2023; Q. Wang et al., 2023; X. Wang et al., 2023). The outcompeting of XBB.1.5 over BQ.1.1 or other co-circulating variants was assumed to be driven by increased ACE2 binding and slightly higher antibody evasion capability of XBB.1.5 (Addetia et al., 2023; Yue et al., 2023). In contrast, the outcompeting of BA.2.86/JN.1 over XBB* variants was mainly contributed to substantially higher immune escape capability together with a slight contribution of increased fitness of the BA.2.86/JN.1 variants (Jeworowski et al., 2024; Planas et al., 2023). Regarding the dominance of JN.1 subvariant KP.3.1.1 and XEC, it has been shown that these two JN.1 subvariants displayed enhanced humoral immune evasion and antibody escape capabilities compared to multiple other co-circulating JN.1 subvariants (Liu et al., 2024).

Regarding specific mutations, XBB.1.5 has been shown to have enhanced infectivity in CaLu-3 cells (Qu et al., 2023) compared to BA.1 or BQ.1.1, which was assumed to be driven by the increased binding of XBB.1.5 to the ACE2 receptor (Yue et al., 2023) owing to the mutation F486P as supported by a structural modeling (Qu et al., 2023). In the early XBB* variants, it was not F486P, but F486S first taking place. F486S in XBB was predicted to cause decreased ACE2 affinity due to the introduction of energetically unfavorable contacts between the polar residue and a hydrophobic patch, but the further mutation of S486P to a great extent reversed this effect by upregulating the propensity of spike protein for hydrophobic interactions with ACE2 and the flexibility of this region, thus allowing improved ACE2 utilization and a corresponding increase in cell-cell fusion and S processing for XBB subvariants with the mutation F486P (Qu et al., 2023; Yue et al., 2023). Interestingly, although most unique mutations in XBB.1.5 were not detected in BA.2.86/JN.1*, F486P occurred in BA.2.86/JN.1* as well, suggesting this specific mutation might have a great impact on the fitness of SARS-CoV-2. In addition to F486P, G339H is another XBB.1.5 unique mutation shared by XBB.1.5 and BA.2.86/JN.1*, but its functional role remains unclear (Cao et al., 2023)

In comparison with XBB.1.5*, BA.2.86/JN.1* bears around 30 different spike mutations. Among these mutations, P681R might be able to improve spike-mediated virus-cell membrane fusion, while K356T, N450D, L452W, A445H, E484K, and V483del are related to increased antibody resistance compared with XBB.1.5 (Yang et al., 2023). In the JN.1 subvariants, the unique mutation S31del (in KP.3.1.1) and T22N (in XEC) have been shown to be able to increase antibody escape from receptor-binding domain-targeting antibodies (Liu et al., 2024). In addition, it has been reported that BA.2.86 was antigenically distinct from BA.5, BA.2, and XBB.1.5 (Yang et al., 2023), which is consistent with the results of phylogenic distance evaluation in the current study. Since the BA.2.86/JN.1* variants are phylogenetically highly distant away from other Omicron variants, their pathogenicity can be different as well, which need be addressed by further clinical data.

### 4.2 Prediction of future evolution trend of SARS-CoV-2

Furthermore, many efforts have also been made to predict the future evolution trend of SARS-CoV-2. As most unique mutations in XBB* were not detected in BA.2.86/JN.1*, this means evolutionally the possibility of that BA.2.86* was a descendent of XBB* is low (Planas et al., 2023), although XBB* had been the dominant variants for more than one year before the occurrence of BA.2.86*. How did this happen? Source of BA.2.86 remains unclear. Likely this is the results of evolution pressure through long time co-existing of one BA.2 variant with host (e.g. one immunocompromised person as host). Also, we could not exclude the possibility that new variants may occur in future as the results of further evolution of the XBB* variants. Generally speaking, there are two possible directions for further evolution: **1)**. In the existing mutation positions that have been tested after long duration circulation and evolution, novel AA mutation with higher fitness (for transmission or immune escape) will emerge. **2)**. New mutation positions emerge: this would be the most unpredictable conditions (transmission ability unknow, pathogenicity unknown)

Since both BA.2.86/JN.1* and XBB* are already pretty much a collection of most previously detected mutations in VOCs, the first type of evolution might be not the major pattern of evolution, and very likely the direction of further evolution is the type 2 evolution – emergence of new mutation positions. Regarding this topic, many wet labs have tested certain possibilities. For example, one study carrying out mutation scans of XBB.1.5 and BQ.1.1 reported an ongoing epistatic drift during SARS-CoV-2 evolution (Taylor & Starr, 2023). Based on currently available evidence, what can also be predicted is: the major trend will be increased transmission ability or immune escape ability, instead of increased pathogenicity.

### 4.3. The decisive factor for one new variant getting growth advantage over other co-circulating variants

Also, what we know is, one evolutional jump does not necessarily immediately lead to the emergence of a new dominant variant. We see the cases for both XBB and BA.2.86. The variant with one evolutional jump, despite naturally born stronger immune escape ability, might have no specific growth advantage. However, this new variant can provide more opportunities for further evolution. For example, the original XBB did not display advantage in transmission, but with the further evolution of it, XBB.1.5, XBB.1.6 gained stronger transmission abilities being able to outcompete BQ.1.1 and other co-circulating variants globally. A similar scenario also occurred for BA.2.86. As a further evolutional product of BA.2.86, JN.1 displayed a growth advantage over the XBB* variants (Kaku et al., 2024).

For a new variant with a lot of newly occurring mutations, many positions can escape structure limitations and gain opportunity for evolution, which may increase the chance of new variant occurring with higher fitness or growth advantage. It was expected that along with the evolution course of SARA-CoV-2, the virus would display a trend of pathogenicity decrease. However, the selection factor of antibody escape does not necessarily favor new variant with lower pathogenicity, such as for the case of the Delta variant, which had both increased infection ability and pathogenicity. Therefore, close monitoring the evolution of SARS-CoV-2 is a necessary precaution measure for public health. Even if global spreading of new variant cannot be avoided, precaution measures can be taken to reduce mortality and related economic loss.

## Data Availability

All data produced in the present work are contained in the manuscript

## Acknowledgements

We thank all researchers who are working around the clock to generate and share genome data on GISAID (http://www.gisaid.org). We thank the local testing labs in Saxony for their support for collecting sequencing samples. We thank the Dresden-concept Genome Centre for their sequencing efforts.

## Financial support

B.Y. is in part supported by a funding from German Research Foundation (DFG Project Number: 458912928; DA 592/12-1 | YI 175/1-1).

## Competing interests

The authors declare none.

## Code availability

Data processing and visualization was performed using publicly available software, primarily RStudio v1.3.1093. Code for constructing phylogenetic maximum likelihood (ML) and time trees as well as phylogeographic analyses is available at https://github.com/genomesurveillance/delta-variant-sublineage, which is modified from SARS-CoV-2-specific procedures github.com/nextstrain/ncov.

## Notes

### Competing Interest Statement

The authors have declared no competing interest.

### Summary of Updates

author affiliation updated, with a new home institute being added.

